# The Medical Knowledge Oligarchies

**DOI:** 10.1101/2023.06.02.23290881

**Authors:** João Matos, Luis Nakayama, Marie-Laure Charpignon, Jack Gallifant, Mohammad Kashkooli, Francesco Carli, Leo Anthony Celi

## Abstract

**Introduction:** Healthcare policies and clinical decisions heavily rely on research publications from high-impact medical journals. A lack of author diversity in medical publications poses a risk to underrepresented groups. To promote equity in healthcare medical decisions, fostering collaborations within research groups is crucial. This study integrates scientometrics with network analysis to uncover intricate co-authorship networks and examine diversity and inclusion in scientific collaboration.

**Methods:** The authors’ metadata from five high-impact medical journals were collected, and a weighted graph of co-authorships was constructed. The study addresses four research questions: identifying influential authors, exploring research output communities, analyzing collaboration patterns, and examining the evolution of collaboration over time.

**Results:** Central nodes are significantly more likely to be male or from high-income countries. Further, when evaluated over time, the graph reveals concerning trends in diversity where collaboration with authors from lower income countries is not growing. All code is publicly available on GitHub.

**Discussion:** The findings underscore the need to promote diversity within research niches and question the role of gatekeepers in facilitating inclusivity. Future studies should expand the scope of network analysis and explore additional factors such as funding sources and guidelines.

**Conclusion:** Overall, this study contributes a framework for auditing diversity and inclusion in scientific collaboration, aiming to promote transparency and a more equitable medical knowledge production system.

## Introduction

Healthcare policies and clinical decisions heavily rely on research publications originating from peer-reviewed medical journals with high impact factors. These journals play a crucial role in evidence-based medicine as they provide essential articles for establishing healthcare policies [1,2]. However, the lack of gender and demographic diversity in medical scientific publications hinders fairness and equal outcomes in healthcare. To address this issue, fostering collaboration and diverse research groups is crucial in reducing existing disparities and the global burden of diseases, paving the way for innovative and creative solutions [3–5].

Scientometrics, as a quantitative study of scientific literature and its attributes, focuses on examining and quantifying scientific activity. Its primary objective is to assess the impact and significance of research publications [6]. Scientometrics and bibliometric analysis primarily describe author metadata characteristics and citations, missing relationships beyond the single study or group aggregates. Collaboration interactions can be intricate, requiring a complex network analysis to gain a profound understanding of the relationships involved.

Network graph analysis is a field of study that explores the relationships and interactions between entities using mathematical and statistical techniques [7]. By examining interconnected objects within a graph, this approach allows for discovering novel patterns and insights. Utilizing network metrics such as degree centrality, betweenness centrality, or modularity, as well as cluster detection techniques like the Louvain method and Infomap, network graph analysis has proven beneficial in diverse healthcare areas, including predicting potential drug indications [8], identifying RNA functions [9], and assisting in clinical decisions based on electronic medical records [10].

This study aims to integrate scientometrics with network analysis to uncover intricate co-authorship networks and provide a comprehensive understanding of the complex knowledge landscape. By examining co-authorship networks and collaboration patterns, we aim to shed light on diversity, inclusion, and the influential authors shaping the scientific discourse [11]. Ultimately, this research aims to contribute to a more equitable and representative scientific community, promoting innovative research and driving impactful advancements in healthcare. Herein, we develop a publicly available framework for conducting network analysis to audit diversity and inclusion in scientific collaboration and examine the top authors and co-authors who publish in high-impact medical journals, focusing on understanding their collaboration patterns. Furthermore, we evaluate the diversity and inclusion within subgroups of authors to identify potential disparities and areas for improvement.

## Materials and Methods

### Data Extraction: Medical Journals and Publications Metadata

This study encompasses the metadata of authors of all research publications from five high-impact medical journals. To gauge the significance and influence of each journal, as measured by the average number of citations received by research articles, we utilized the Journal Citation Report ranking provided by Clarivate Analytics within the “General and Internal Medicine” category [12].

We selected the top four journals retrieved: The Lancet (established in 1823), New England Journal of Medicine (NEJM, established in 1812), Journal of the American Medical Association (JAMA, established in 1883), and British Medical Journal (BMJ, established in 1840). Additionally, we included Nature Medicine (established in 1995), which held the first position in the “Medicine, Research & Experimental” category.

The Dimensions AI platform, a linked research information dataset, was used to collect metadata pertaining to each research article, including every article captured/indexed by DimensionsAI published between January 2007 and December 2022, regardless of their type (e.g., observational studies, RCTs, editorials, systematic reviews, patient case reports) [13].

### Feature Engineering: Authors’ Gender and Country

We cross-linked the Dimensions platform to match the article’s authors and retrieve the existing IDs, name variants, affiliation data, research topics, published journals, co-authors, and active years.

Each author’s first name was then extracted using metadata and processed by the Genderize.io Application Programming Interface (API). The API has a collection of names that have been previously annotated and linked to their reported gender. Given the data stored within, the API can calculate the probability of male or female gender and assign the most probable one to the first name being analyzed.

Each author was also linked to a country based on their affiliation. For authors who had multiple affiliations, all were considered separately. To analyze the representation of low and middle-income countries (LMIC) and high-income countries (HIC), we considered the World Bank Country and Lending Groups dataset, which ranks nations according to their gross national income (World Bank country and lending groups). We cross-linked the Dimensions and World Bank classification, which allowed us to create a new variable characterizing to analyze the proportion of authors from LMICs and HICs.

### Cohort Selection

After extracting the data from Dimensions, and cross-linking with the gender information, we excluded publications with a single author (don’t contribute to the network), publications with more than 100 authors (all authors may not actually know each other), and authors with missing gender information.

### Network Analysis: Weighted Graph of Co-Authorships

We constructed a weighted, undirected graph. Authors are represented as nodes; edges represent co-authorships, and their weights represent the number of publications shared by each pair of authors (see **Figure 2**).

### Research Questions and Approaches

The main question, *“Is scientific collaboration in high-impact medical journals diverse and inclusive enough?”* was further split into four questions and approaches.

### Question 1: Who are the network’s most influential and central authors?

#### Analysis of nodes with the highest centrality metrics

The following centrality measures were computed for each node of the built network:

- *Degree Centrality:* Counts the number of connections a node has in a network, indicating its popularity or importance.
- *Betweenness Centrality:* Measures a node’s position as a bridge between other nodes in a network, highlighting its control over information flow or influence.
- *Closeness Centrality:* Gauges a node’s proximity to others in a network, indicating its potential for efficient information transfer or influence [14].

Subgroups of nodes - according to their centrality percentile - were then assessed in terms of LMIC and female presence. The goal was to answer the question “*What is the LMIC and female representation in the top x % most central nodes”*, where *x* varied between 1-100%. Nodes in such subgroups should represent the most influential authors of the network.

A two-sample T-test was performed at each percentile to assess whether the LMIC and female distributions of each subgroup were statistically different from the complete network. The null hypothesis was rejected at a p-value < 0.05.

### Question 2: Are there communities of authors with high research output?

#### Analysis of top maximal cliques

A clique is a subset of nodes in a graph where every node is directly connected to every other node in the subset; a maximal clique is a clique that cannot be expanded further.

We propose that maximal cliques with a high number of authors and/or publications can be viewed as a representation of influential and significant communities within the network.

### Question 3: Who collaborates with whom, and how often?

#### Analysis of edge types

We analyze the distribution of edge types in the network to determine the diversity of collaboration between authors, considering gender and country. These include the following combinations:

- *Gender:* F-F, F-M, and M-M
- *Country:* L-L, L-H, and H-H
- *Joint:* LF-LF, LM-LM, LM-LF, HF-HF, HM-HM, HM-HF, LF-HF, LM-HM, LM-HF,

where F: Female, M: Male, L: LMIC, and H: HIC.

### Question 4: How has collaboration evolved from 2007 to 2022?

#### Analysis of edge types through the years

To analyze the temporal evolution of the network, we assess the edge types, similarly to question 3, but stratified by year, at different granularities:

- *Yearly:* 2007-2022, where an isolated graph is created for each year’s publications
- *4-Year Bins:* 2007-2010, 2011-2014, 2015-2018, 2019-2022, obtaining one graph per period, resulting in a total of four different graphs.

### Software and Code Availability

All analyses were performed with Python 3.10.9. Data cleaning was done with Pandas [15]; NetworkX [16] and NetworKit [17] packages were used for network analysis; statistical tests were performed with NumPy [18] and SciPy [19]; visualizations with Matplotlib [20]. All code is available on GitHub.

## Results

The search strategy retrieved 93,751 articles and 172,060 authors. After applying our exclusion criteria, we were left with 50,042 articles and 145,878 authors (see **Figure 1**).

**Figure 1.**
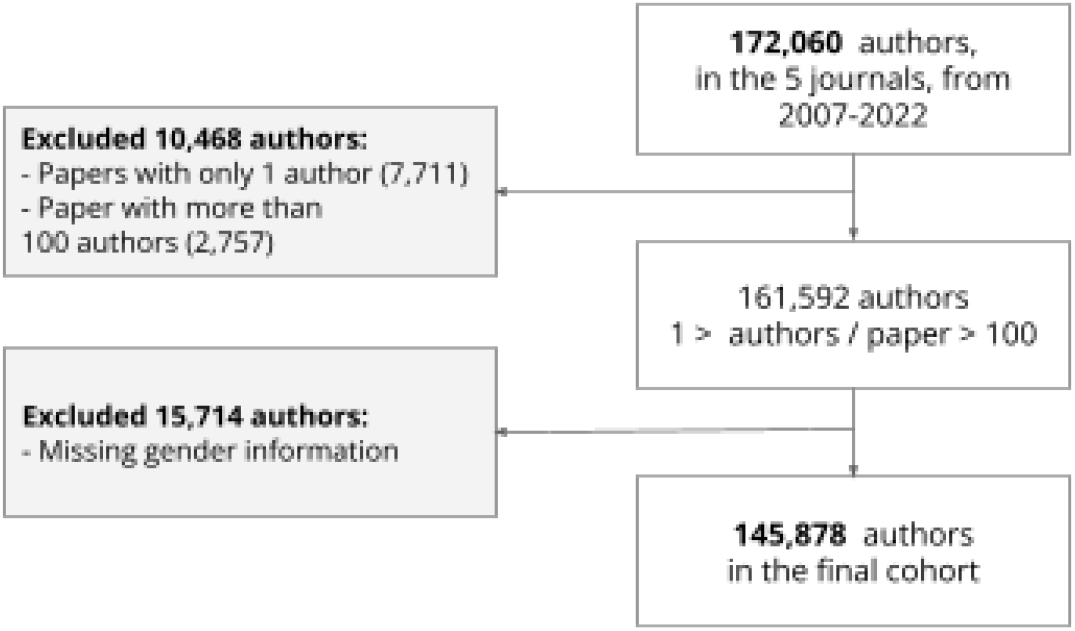
Flow diagram with data preprocessing methodology.

**Figure 2.**
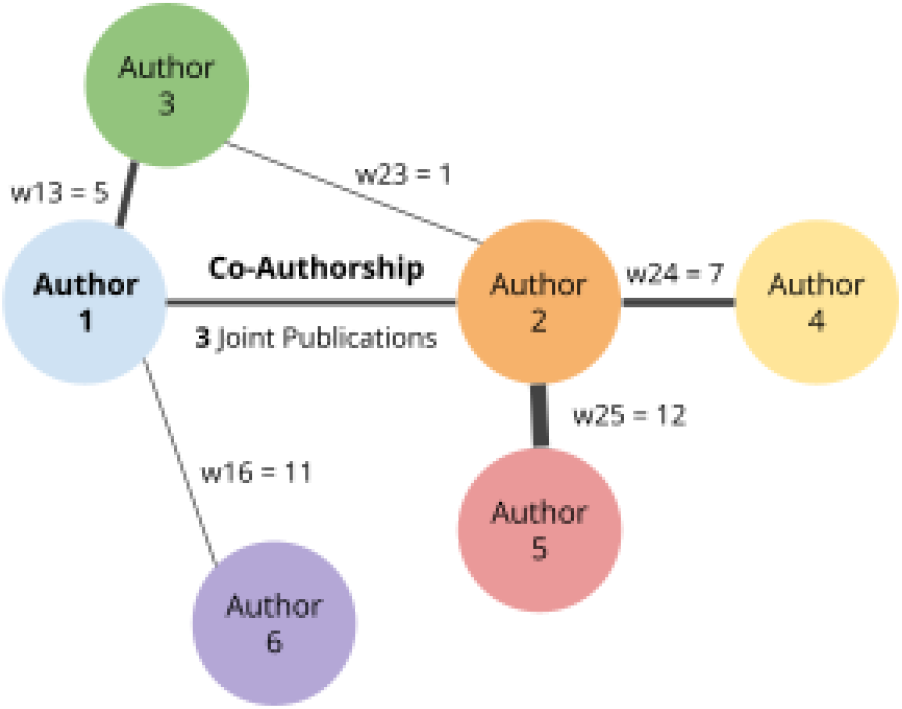
Example of Co-Authorship Weighted Graph. Nodes are the Authors; Edges are a Co-Authorship; Weights are the number of publications together

**Table 1** provides summary statistics of the final cohort. The journal with the most publications was “The Lancet” – 52,848 (36.2%) – and the one with the least was “The BMJ” – 29,525 (20.2%). The journal with the most number of authors was “The Lancet” as well – 16,449 (32.9%) – but, interestingly, “Nature Medicine” presents the least number of authors – 3,883 (7.8%). The overall median [IQR] of authors per publication is 3 [2, 6]; notably, Nature Medicine presents 10 [3, 17] authors per publication. Since 2007, the number of authors and publications has been growing. There are 93,305 (33.3%) female authors and 23,706 (8.5%) authors coming from LMICs.

**Table 1.**
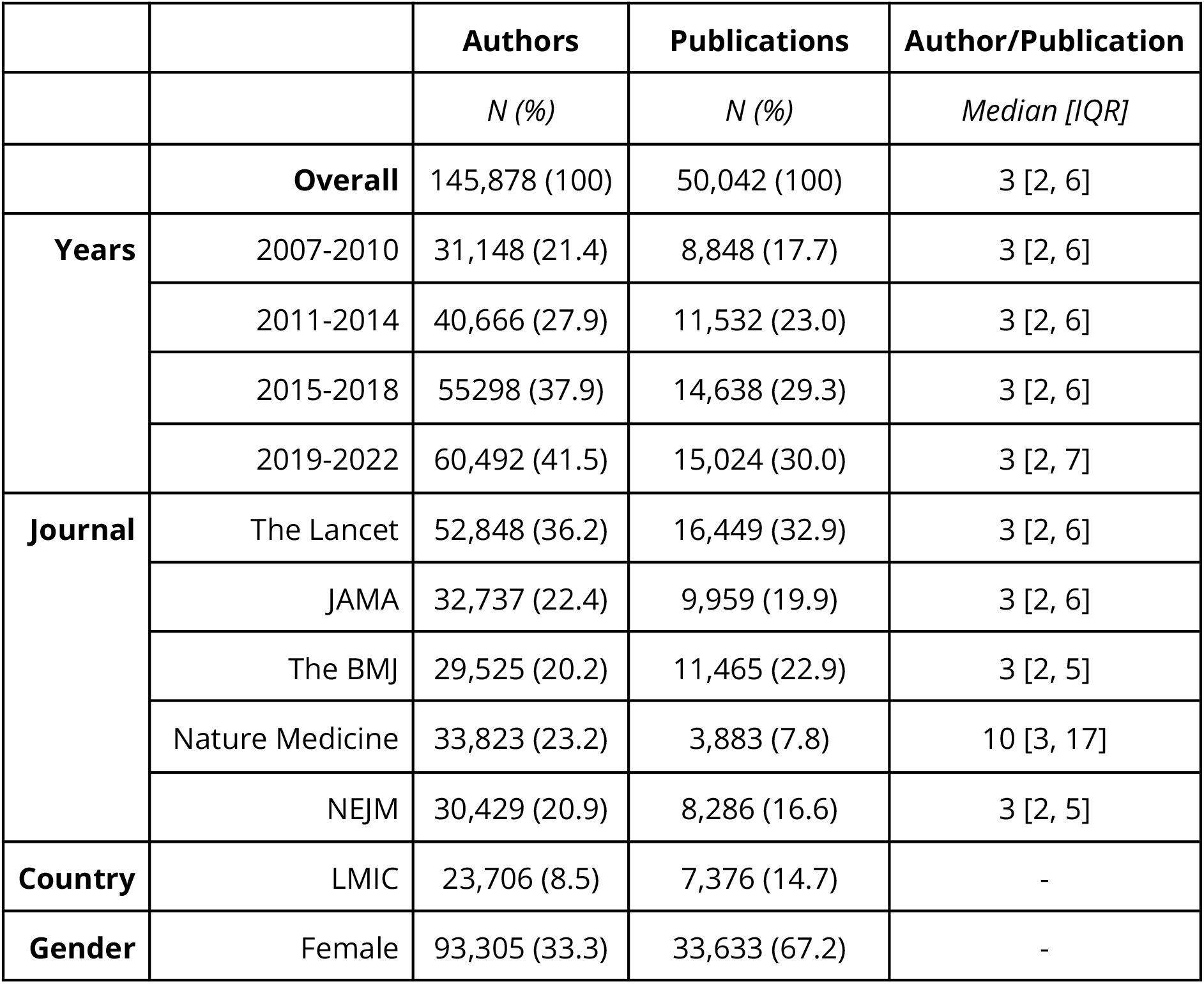
Summary Statistics of the final cohort.

|  |  | **Authors** | **Publications** | **Author/Publication** |
| --- | --- | --- | --- | --- |
|  |  | *N (%)* | *N (%)* | *Median [IQR]* |
|  | **Overall** | 145,878 (100) | 50,042 (100) | 3 [2, 6] |
| **Years** | 2007-2010 | 31,148 (21.4) | 8,848 (17.7) | 3 [2, 6] |
|  | 2011-2014 | 40,666 (27.9) | 11,532 (23.0) | 3 [2, 6] |
|  | 2015-2018 | 55,298 (37.9) | 14,638 (29.3) | 3 [2, 6] |
|  | 2019-2022 | 60,492 (41.5) | 15,024 (30.0) | 3 [2, 7] |
| **Journal** | The Lancet | 52,848 (36.2) | 16,449 (32.9) | 3 [2, 6] |
|  | JAMA | 32,737 (22.4) | 9,959 (19.9) | 3 [2, 6] |
|  | The BMJ | 29,525 (20.2) | 11,465 (22.9) | 3 [2, 5] |
|  | Nature Medicine | 33,823 (23.2) | 3,883 (7.8) | 10 [3, 17] |
|  | NEJM | 30,429 (20.9) | 8,286 (16.6) | 3 [2, 5] |
| **Country** | LMIC | 23,706 (8.5) | 7,376 (14.7) | - |
| **Gender** | Female | 93,305 (33.3) | 33,633 (67.2) | - |

**Table 2** provides a description of the produced network for 2007-2022. Interestingly, the 145,878 nodes (authors) produce 1,352,110 edges (pairs of co-authors publishing at least once together). The average clustering coefficient was 79.7 % and there were 6,255 connected components.

**Table 2.**
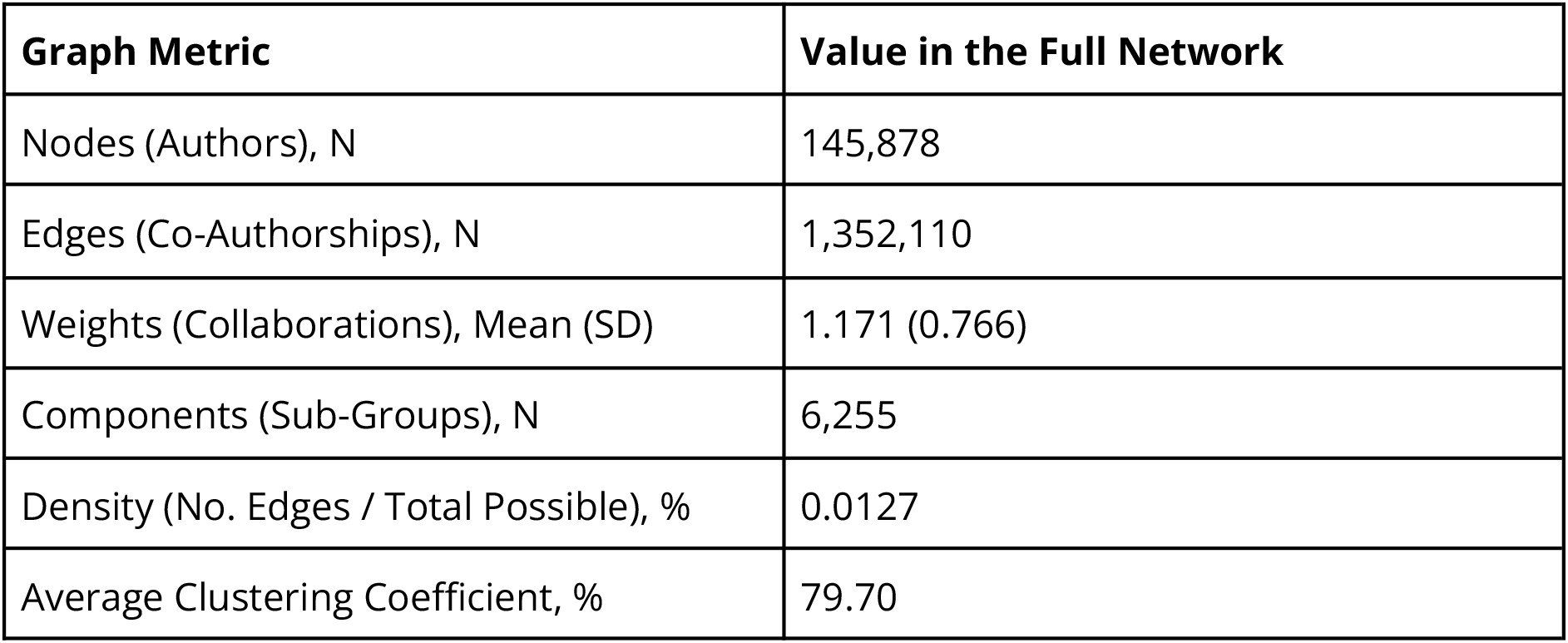
Summary Statistics of the Weighted Graph.

| Graph Metric | Value in the Full Network |
| --- | --- |
| Nodes (Authors), N | 145,878 |
| Edges (Co-Authorships), N | 1,352,110 |
| Weights (Collaborations), Mean (SD) | 1.171 (0.766) |
| Components (Sub-Groups), N | 6,255 |
| Density (No. Edges / Total Possible), % | 0.0127 |
| Average Clustering Coefficient, % | 79.70 |

### Question 1: Who are the network’s most influential and central authors?

The more central a node is, the less likely it is to be an LMIC or female author (see **Figures 3a, 3b**, and **3c**). For example, among nodes with the top 1% betweenness centrality, only 6% are from LMICs (vs. 8.5% in the whole network), and 15 % are female (vs. 33.3% in the whole network). Statistically significant differences were consistently found for all metrics, at least, from percentile 1% to 55%.

**Figure 3a.**
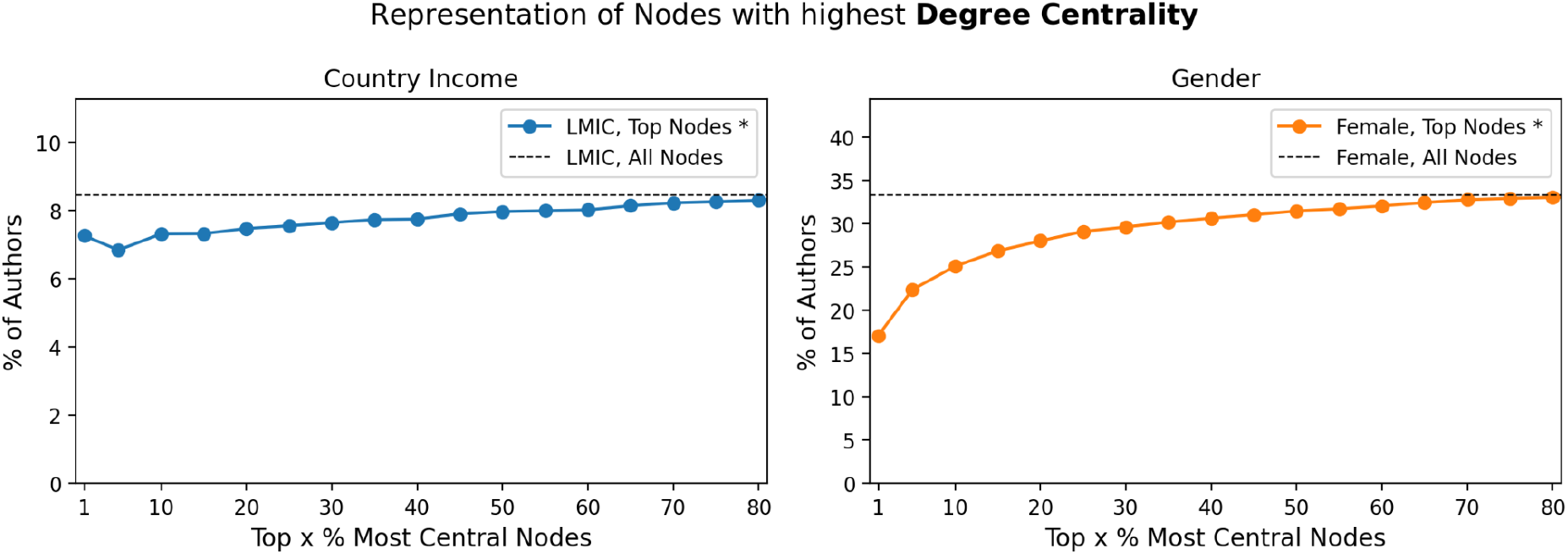
Country income and Gender of authors with highest <u>degree centrality</u>.

**Figure 3b.**
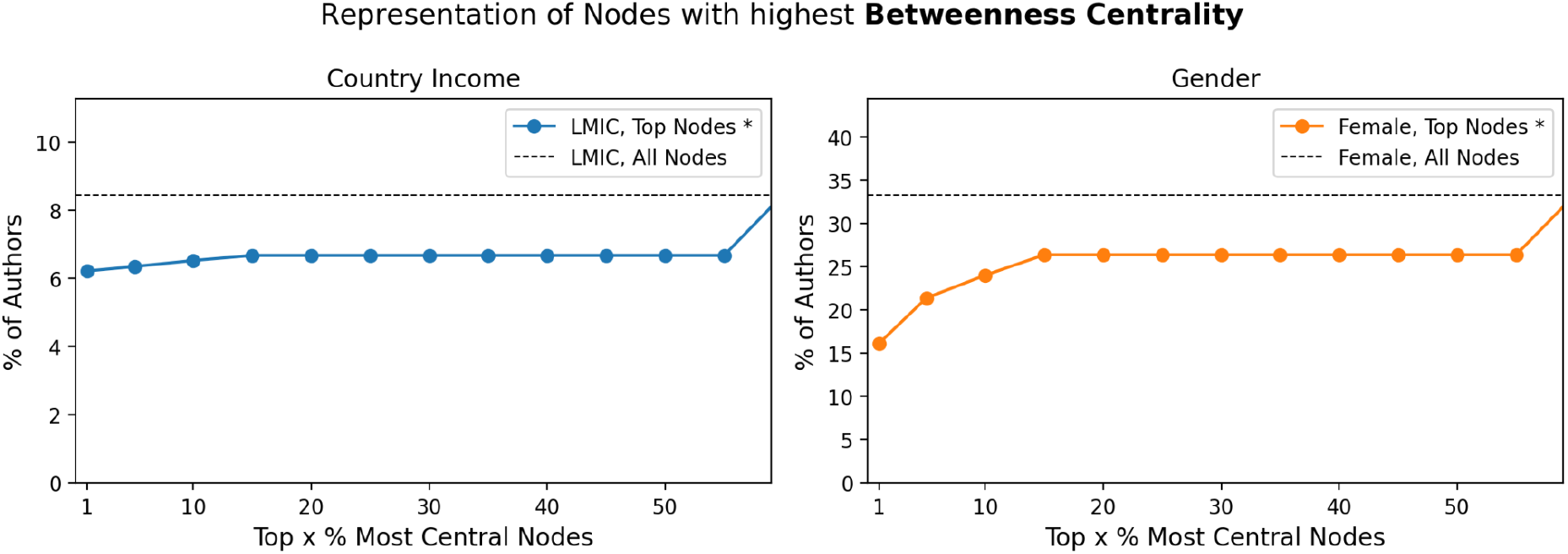
Country income and Gender of authors with highest <u>betweenness centrality</u>.

**Figure 3c.**
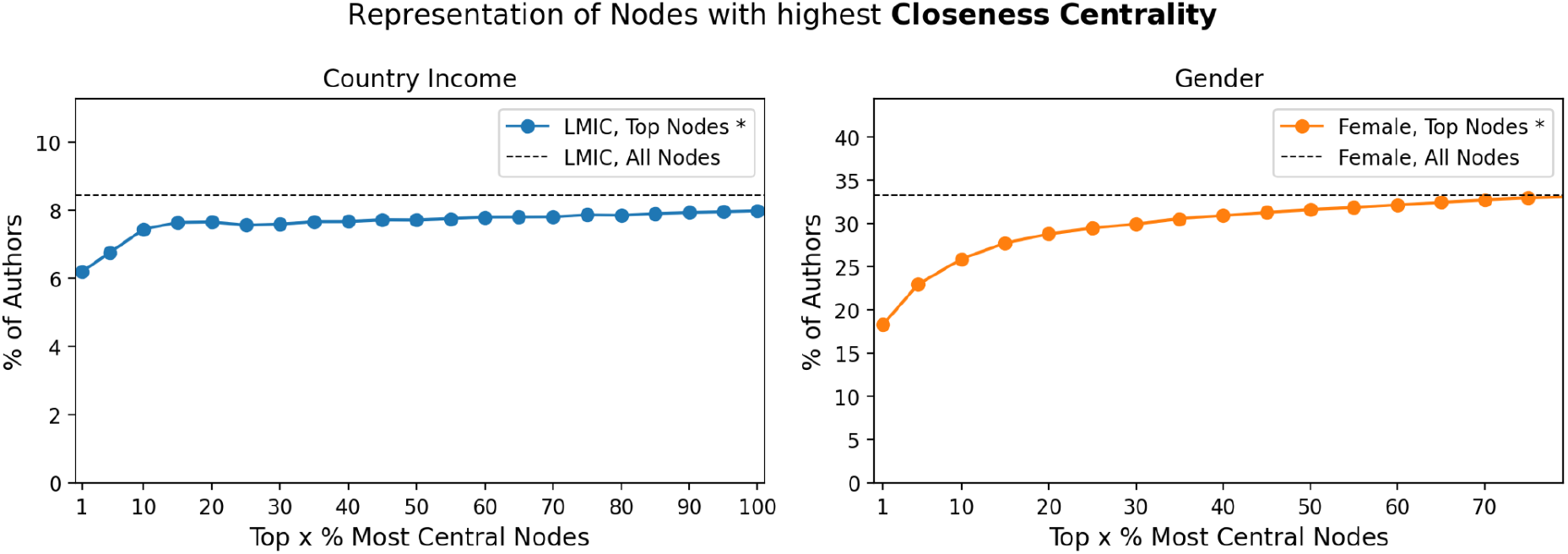
Country income and Gender of authors with highest <u>closeness centrality</u>. Displayed values present a p-value < 0.05, after performing a two sample T-test between the set with all nodes and the subset with most central nodes, at each top x %. As we go from the left to the right in the x axis, we get away from most central nodes.

These findings highlight the lack of diversity and inclusion of minority authors in nodes with more popularity and importance within the network. This trend is consistent across the different centrality measures (degree, betweenness, and closeness).

### Question 2: Are there communities of authors with high research output?

**Figures 4a** and **4b** depict the composition of the top 5 maximal cliques with a maximum number of publications and authors, respectively. All these cliques have more HIC and male authors than the network overall, showcasing a lack of inclusivity to some extent.

**Figure 4a.**
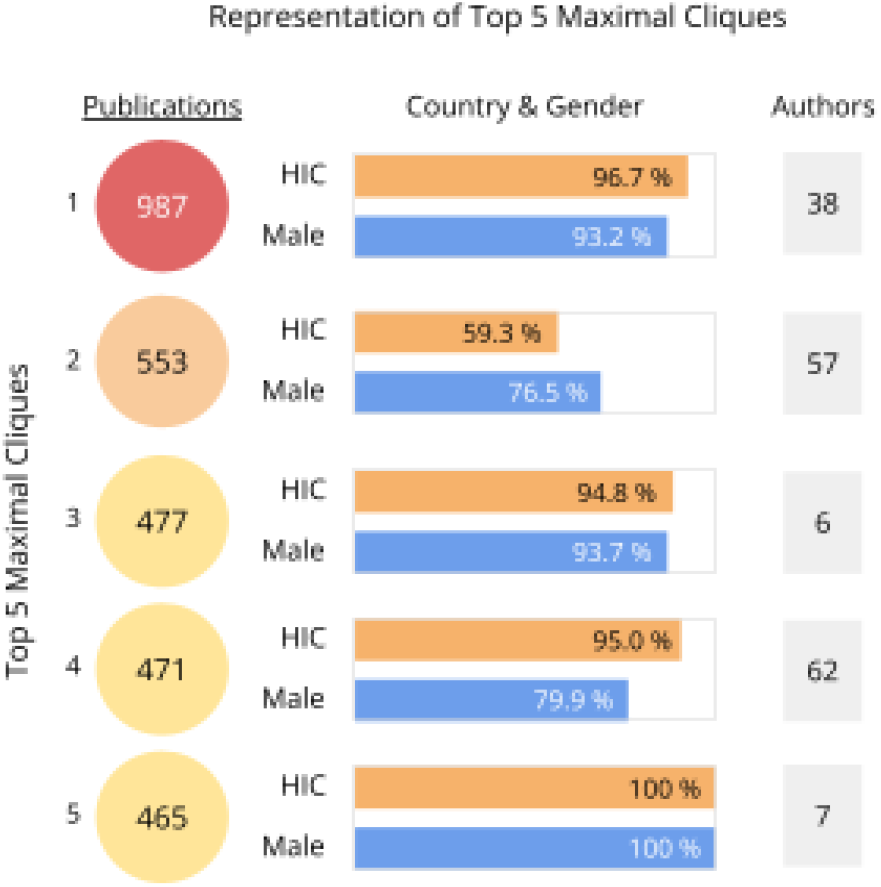
Representation of top 5 maximal cliques with most publications.

**Figure 4b.**
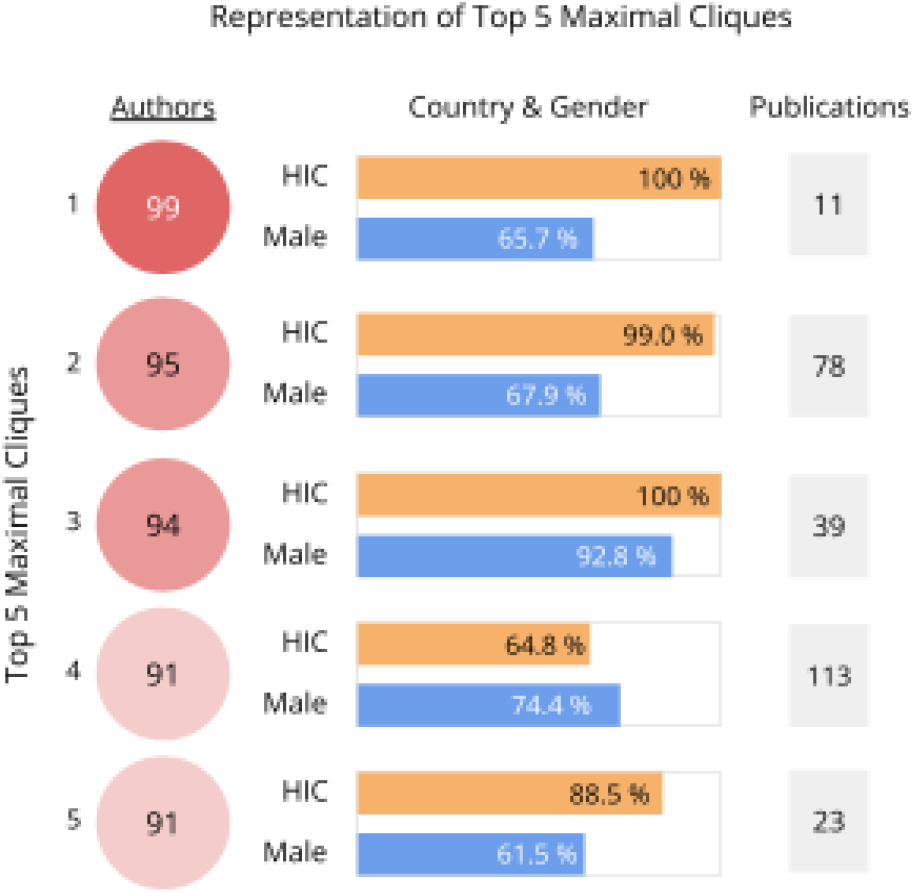
Representation of top 5 maximal cliques with most authors, from 2007-2022.

We found that the co-authorship network does contain communities with strong scientific output. For example, the clique with the most publications, with just 38 authors (93.2% male and 96.7% from HIC) produced 987 publications alone. Another clique, with just 7 authors, 100% male and 100% from HIC, produced 465 publications.

Further investigation is needed to truly understand who these authors are and why they present such research output. Furthermore, there are no maximal cliques with more than 99 authors, which is the limit that we set for authors per publication, suggesting that these cliques may be limited to a single publication.

### Question 3: Who collaborates with whom, and how often?

**Figure 5** presents the distribution of edge types. As expected, Male-Male (43.6%) and HIC-HIC (88.1%) edges are predominant, but interestingly, Female-Male edges present a comparable representation, with 42.6%. This number is particularly high when compared to Female-Female edges – 13.8% – showing either more openness to collaboration or inter-dependency between male and female authors, an interesting finding given that female authors are a clear minority in the network.

**Figure 5.**
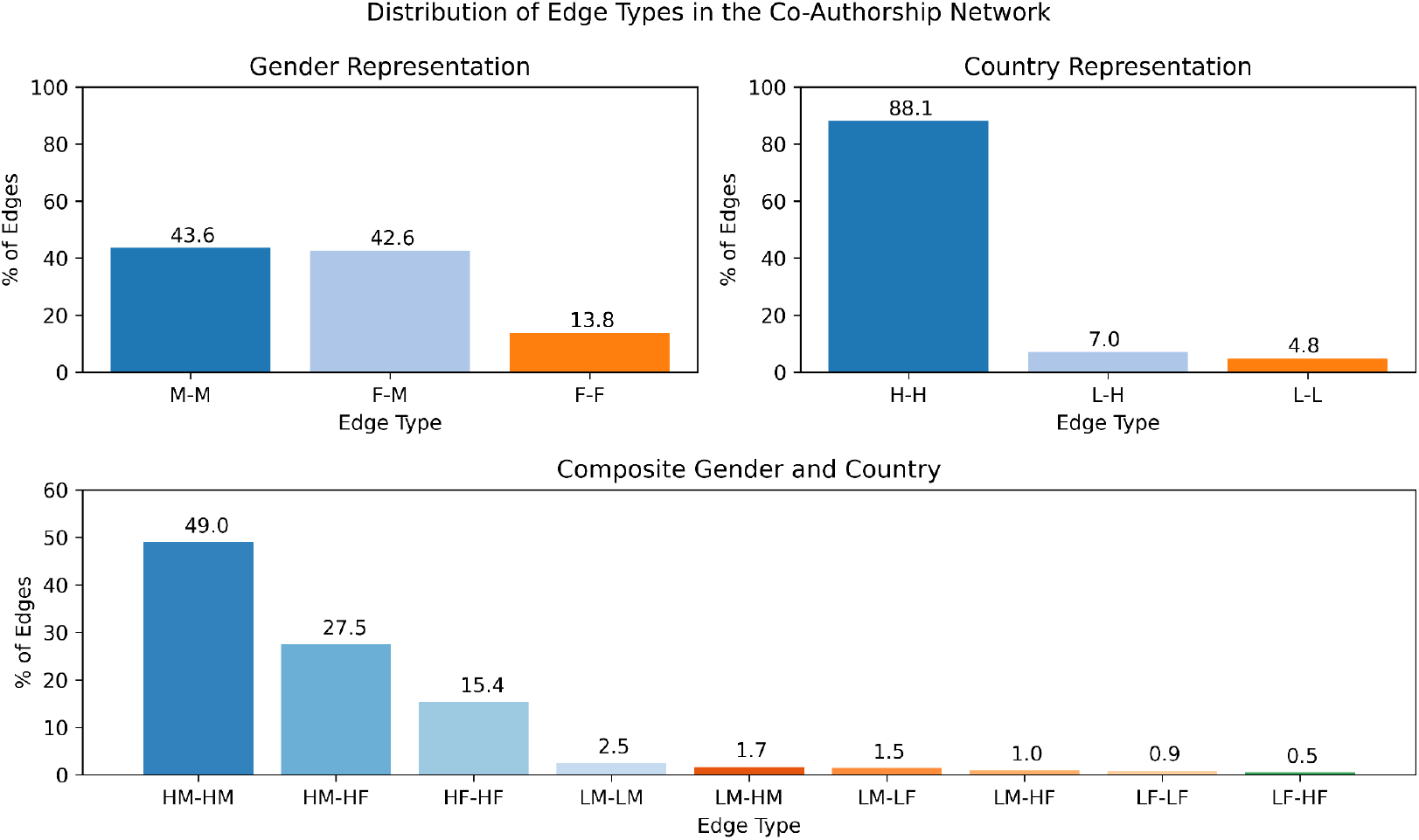
Distribution of Edge Types, encompassing gender, country income, and interactions of both.

Similarly, LMIC-HIC represents 7% of the edges, while LMIC-LMIC 4.8%, which could suggest some dependency of LMIC authors on HIC authors to publish. However, further investigation is needed to ascertain the direction of such dependency.

When looking at the combination of gender and country in the edges, interesting patterns arise as well. Notably, authors with the same gender coming from LMICs are more likely to collaborate with LMIC authors than HIC authors (LM-LM: 2.5% vs. LM-HM: 1.7%; LF-LF: 0.9% vs. LF-HF: 0.5%).

### Question 4: How has collaboration evolved from 2007 to 2022?

**Figure 6** shows the evolution of yearly networks, where there is a growing no. authors, publications, co-authorships, co-authorships per author, authors per publication, LMIC, and female representation.

**Figure 6.**
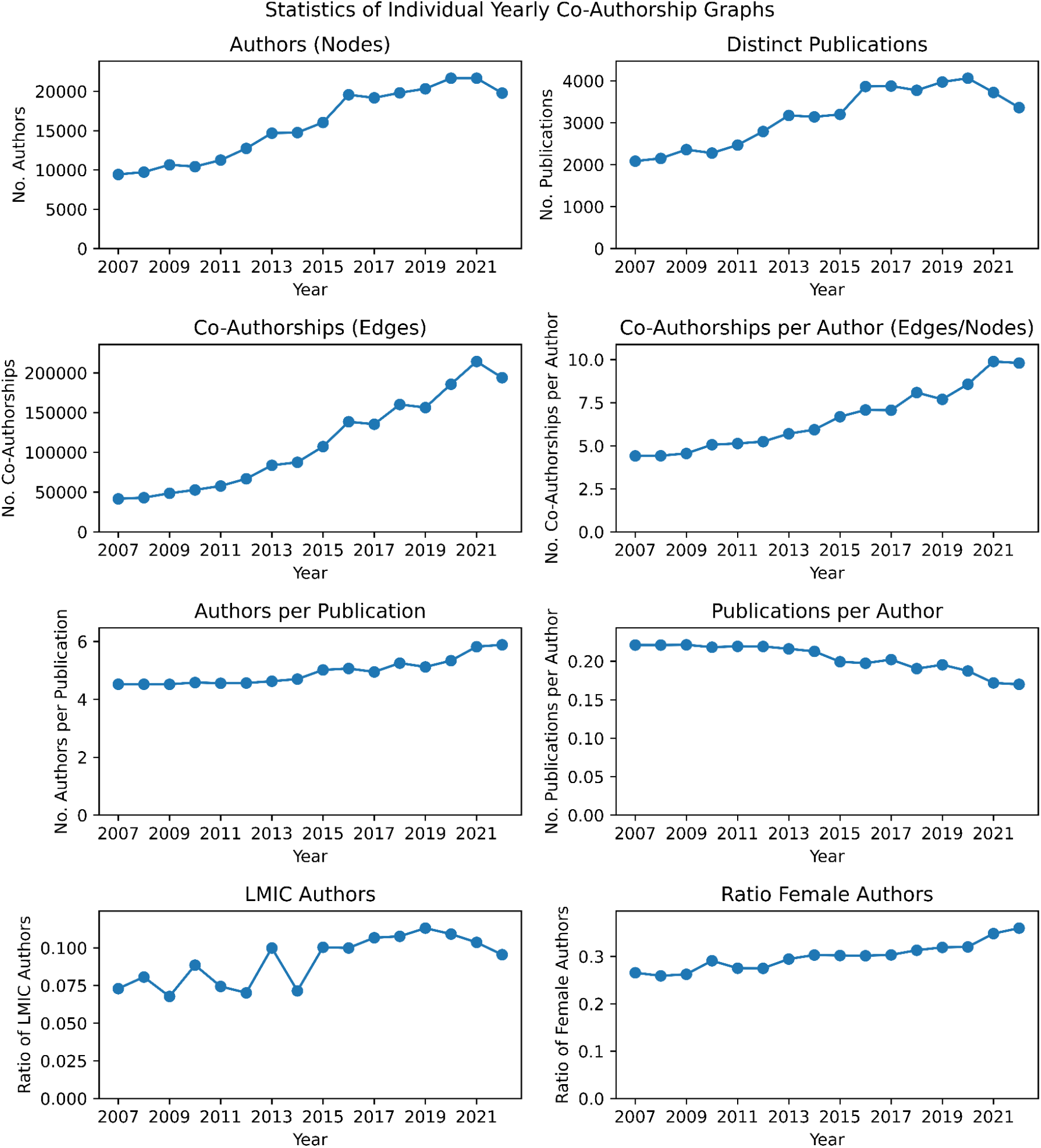
Summary statistics of yearly co-authorship graphs.

**Figure 7a** depicts the change in distribution of edge types with respect to country income, from 2007 to 2022. Despite small variations, unfortunately, there seems to be no significant change in collaboration between LMIC and HIC authors.

**Figure 7a.**
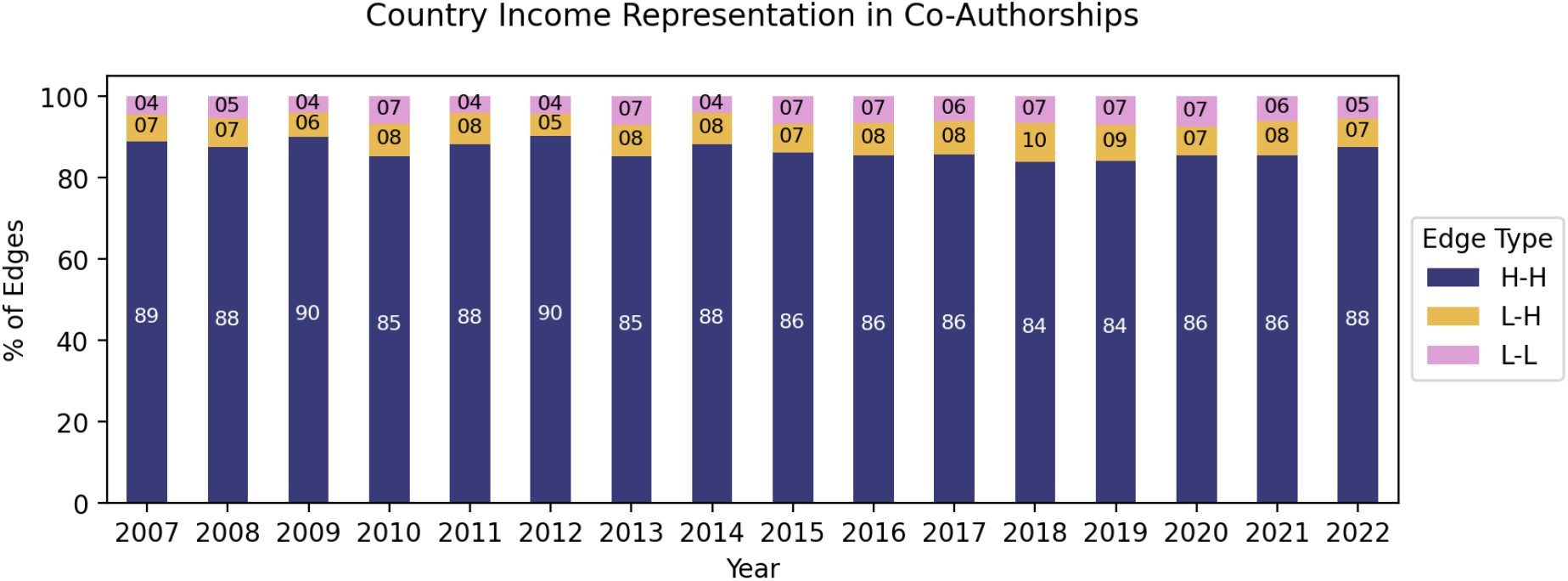
Diversity of co-authorships per year, in terms of authors’ country income. **Abbreviations:** H, High Income Country (HIC); Low- to Middle- Income Country (LMIC)

**Figure 7b** reports the evolution of edge types representation in terms of gender: as the number of female authors is growing, so is the amount of edges of type F-M (37% in 2007 vs. 44 in 2022) and F-F (9% in 2007 vs. 14% in 2022).

**Figure 7b.**
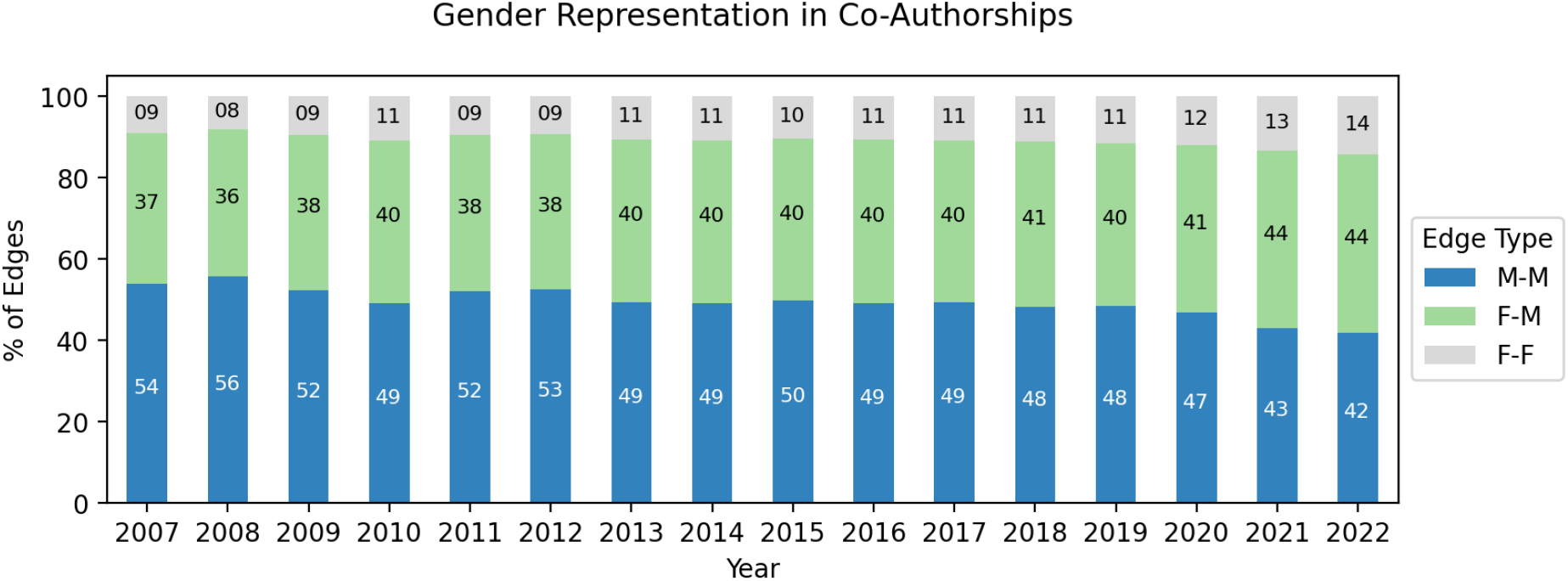
Diversity of co-authorships per year, in terms of authors’ gender. **Abbreviations:** M, Male; F, Female

**Figure 8** shows similar trends for the joint edge types: in general, since 2007, collaboration among minorities has been increasing, showing an improvement in the *status quo*.

**Figure 8.**
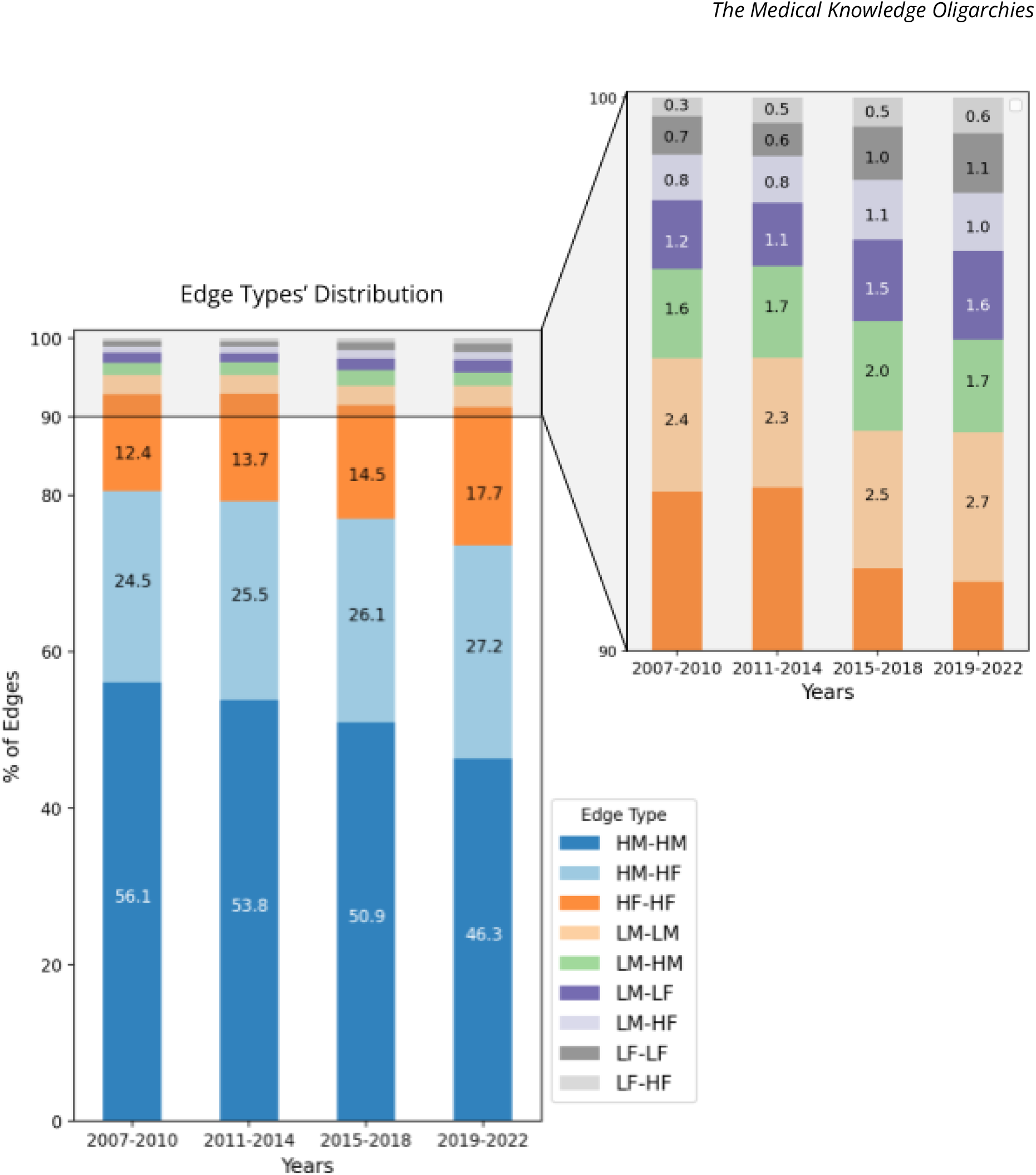
Joint representation of co-authorships’ gender and country, every 4 years. **Abbreviations:** H, High-Income Country (HIC); L, Low- or Middle-Income Country (LMIC); M, male; F, female

### A Framework to Audit Diversity and Inclusion in Scientific Collaboration

More than the numbers and specific findings for this knowledge domain, the main result of this study is the set of methodologies that we share with the community. All code is openly available here: https://github.com/joamats/mit-knowledge-oligarchies.

## Discussion

In this study, we have revealed critical insights into research authorship through the lens of network analysis. One of the key findings is the capacity to identify cliques or interconnected social and professional networks within the publications in high-impact medical journals [11]. By mapping these networks over time, we are able to discern the impact of co-authorship and its changing dynamics. This paves the way for further research into the causal influence of co-authorship with a specific individual, group, or institution on future citations and niche dominance. However, our results display some concerning trends when analyzing diversity within these research niches. Individuals with higher importance and centrality within their respective niches are more likely to be male and from HIC. At the same time, they are less likely to be female or from LMIC. This observation is consistent with prior scientometric analyses, but the use of network analysis offers a more comprehensive understanding of how co-authorship fits within the broader research landscape [21–23].

We also found that diversity within specific research niches is gradually improving for female representation, but not so much for lower income countries. This raises the question of whether the gatekeepers of these established niches are facilitating or impeding progress toward greater diversity.

Nonetheless, our study has limitations. For instance, identifying individuals within the network can be challenging, and there is a clear need for more rigorous causal inference methods to allow for robust hypothesis testing. Moreover, our analysis was limited to only five journals and the last sixteen years, which may not represent the full spectrum of research authorship. Finally, as we are only assessing authors who actually published in these high-impact journals, the minorities of authors that we are looking at may be poorly labeled and not accurately represent the ones who are marginalized.

Future studies should seek to expand the scope of network analysis in research authorship. This includes investigating various fields and exploring different types of co-author demographics. We also aim to link these findings to other aspects of the research ecosystem, such as funding sources, guidelines, dataset characteristics, and citations. In doing so, we believe that this powerful network analysis tool can shed light on even more nuanced and complex dynamics within the world of research authorship.

## Conclusion

The scope of this study goes beyond the findings and numbers we report. We provide a set of tools and methodologies to thoroughly examine who publishes with whom, who is being marginalized from scientific collaboration, and who is controlling medical knowledge production. Ultimately, we propose and make available a scientometrics framework based on network analysis that we hope to promote further studies.

As we advocate that medical knowledge must not be produced by a limited group of researchers – but by a broad, diverse, and inclusive community – we hope that this work is a step towards more transparency in the process of scientific publication.

## Data Availability

All code and data produced are available online at https://github.com/joamats/mit-knowledge-oligarchies

